## Supplementary material for "Interactions between climate change, urban infrastructure and mobility are driving dengue emergence in Vietnam": Electronic supplementary material document

**Supp. Figure 1: District-level dengue timeseries across a latitudinal gradient of transmission settings in Vietnam.** Heatmap shows the full district-level dataset of monthly dengue cases from May 1998 to April 2021 (n=174,936 observations), with colour scale showing natural log-transformed monthly crude incidence (cases per 100,000 inhabitants). Panels show dengue incidence timeseries at district-level (n=667 total with each row representing a district) for each of the eight climatic subregions of Vietnam, which are ordered by latitude from highest (Northeast) to lowest (Mekong River Delta). Within each subregion, districts are ordered by latitude from highest to lowest. Cells shaded grey are missing observations and show when the time series data commence (from 1998 in the north, and between 1999 and 2001 in the south). Vietnam's five major urban municipalities are in the Red River Delta (Ha Noi, Hai Phong), South Central Coast (Da Nang), Southeast (TP. Ho Chi Minh) and Mekong River Delta (Can Tho).

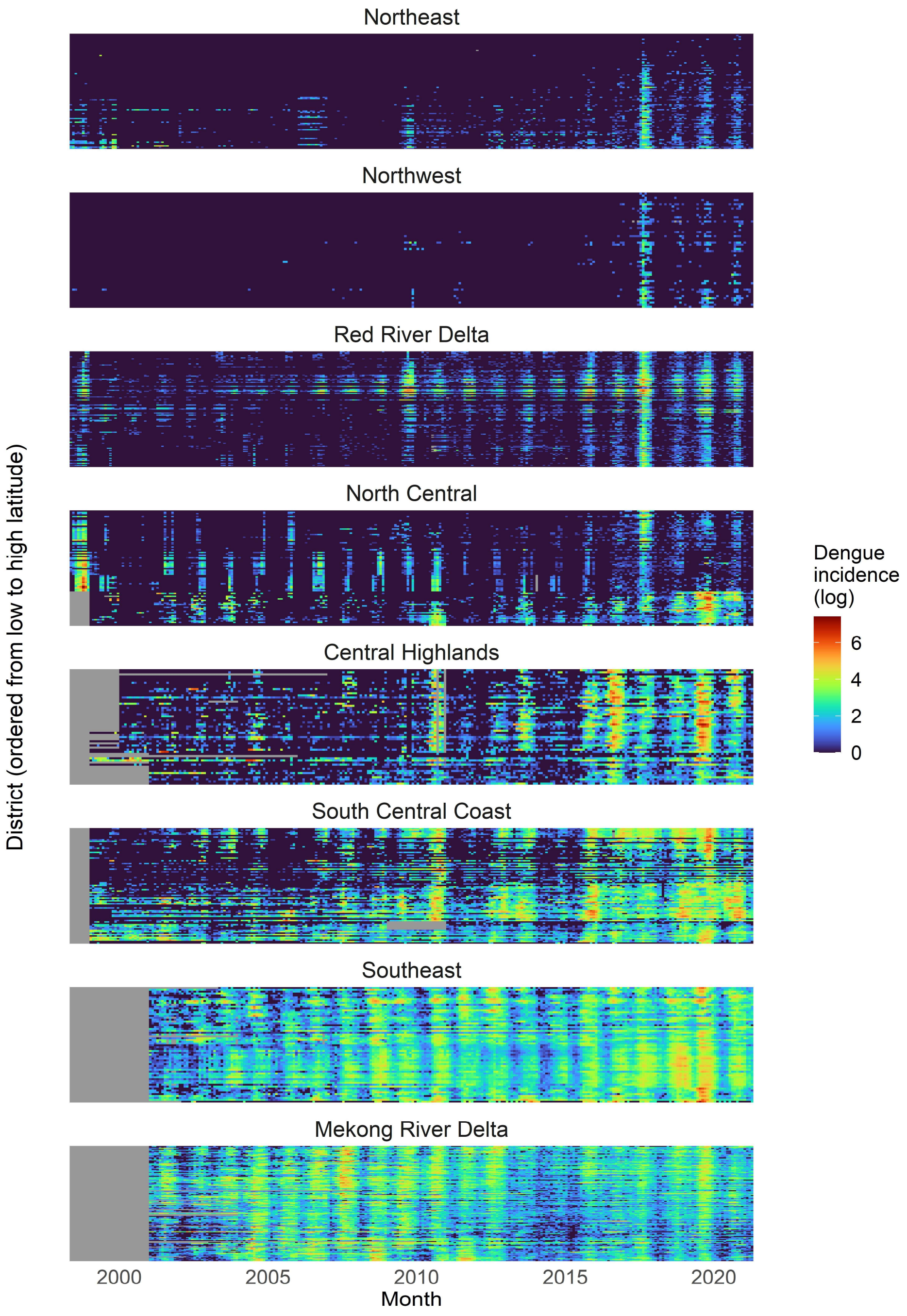

**Supp. Figure 2: Geographical distribution and correlation matrix of socio-environmental covariates across Vietnam.** Maps show the geographical distribution of annual district-level socio-environmental covariates in 2009 (the middle of the study period): log population density, built-up land proportion, mean urban expansion rate in preceding 3 and 10 year windows ($km^2$ per year), access to piped or drilled well water (proportion households), access to flush toilet (proportion households), log gravity and radiation fluxes, road travel per inhabitant (km per person per year), and $T_{mean}$ of the coolest month (°C). The matrix plot shows pairwise correlation coefficients between all socio-environmental covariates for the full dataset (i.e. across all years 1998 to 2020), excluding self-to-self comparisons, with larger point size and darker colour denoting stronger correlation. Very few pairs are strongly correlated ($\rho > 0.7$): population density with built-up land and gravity flux; and urban expansion at short (3 year) and long (10 year) timescales.

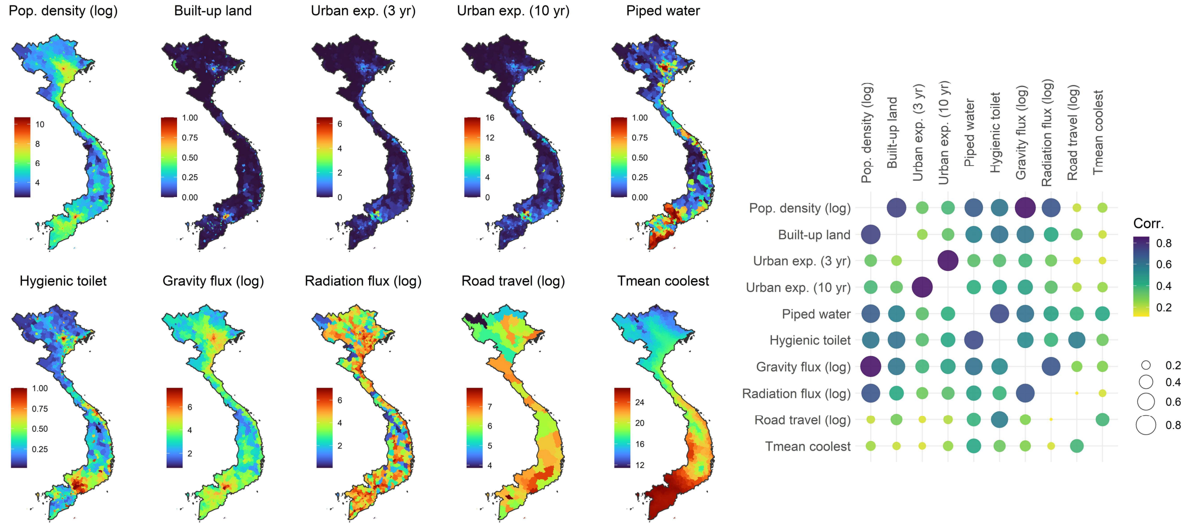

**Supp. Figure 3: Temperature and hydrometeorological dynamics across climatic regions of Vietnam.** Line graphs show district-level monthly mean observations of $T_{mean}$ (daily mean temperature) and precipitation (millimetres per day), and Standardised Precipitation Evapotranspiration Index within 1-month (SPEI-1) and 6-month (SPEI-6) windows. Rows are for different geographical regions of Vietnam, ordered by latitude from north to south ("South" contains both Southeast and Mekong River Delta). Each coloured line shows individual time series for each district, with the black line showing the mean across all districts, i.e. a wider colour band reflects wider intra-regional variation in a climate metric in any given month. SPEI-1 and SPEI-6 measure, respectively, short-term and long-term water excess (values above 0) or deficit (values below 0) relative to the historical average for the same period of the year. Temperature data are derived from ERA5-Land reanalysis, and precipitation and SPEI are derived from WFDE5 and ERA5-Land reanalysis datasets (Methods, Supp. Text 1, Supp. Figure 4).

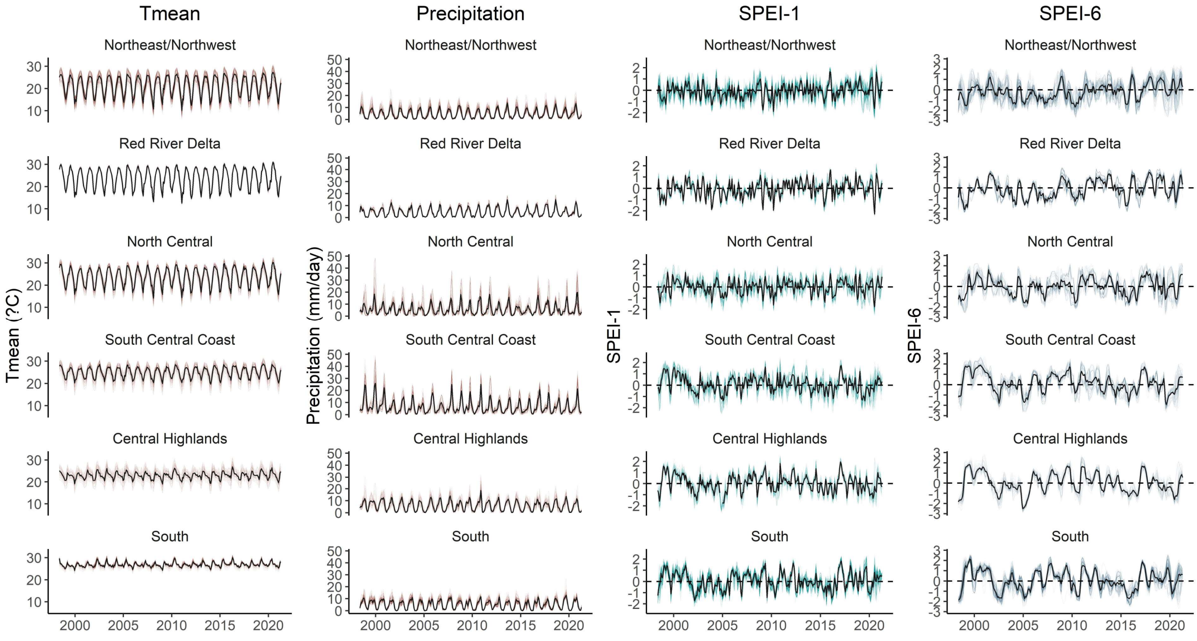

**Supp. Figure 4: Comparison of precipitation observations between weather stations and climate reanalysis products.** Time-series data on monthly precipitation (mm per month) from 15 weather stations across Vietnam between 2002 and 2019 (locations shown in A, source: Vietnam General Statistics Office [https://www.gso.gov.vn/en/](https://www.gso.gov.vn/en/)) were compared to time-series calculated from ERA5-Land and WFDE5 (bias-corrected ERA5) gridded reanalysis datasets (Methods). Bar graph shows, for the whole of Vietnam and for each individual station (ordered by latitude from north to south), a comparison of error between the two reanalysis datasets, measured as the mean absolute difference between all station observations and reanalysis-derived estimates (B). Error was consistently much lower for the bias-corrected WFDE5 dataset than ERA5-Land, so this dataset was used to derive precipitation and SPEI metrics in this study (Methods). Comparisons showing correspondence between monthly station observations (grey bars) and WFDE5-based precipitation estimates (green line) are shown for 6 example stations (C).

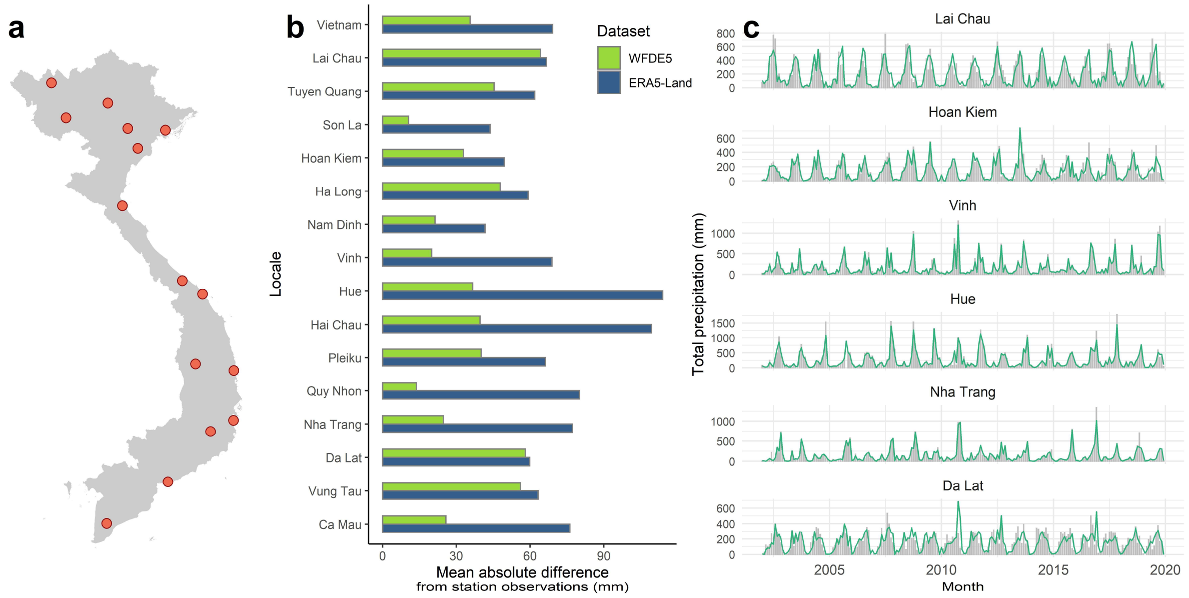

**Supp. Figure 5: Fitted spatiotemporal and seasonal effects from a baseline model of dengue incidence.** The baseline (random effects-only) model of dengue incidence included an offset for population, a province-specific effect of calendar month to account for seasonality (modelled as a first-order random walk) and district- and dengue-year specific spatially-structured and unstructured random effects (Methods). Effects of month for each province (n=63), ordered by latitude from north to south, are shown in (A), where polygon height denotes dengue relative risk and polygon colour denotes geographical region (as shown in map B). Mean fitted district-level effects (structured + unstructured, averaged across all years) are shown in (C), where the colour scale denotes contribution to expected log dengue incidence, from lowest (dark green) to highest (dark brown). Dengue transmission intensity decreases, and becomes more strongly seasonal, along the south-to-north latitudinal gradient.

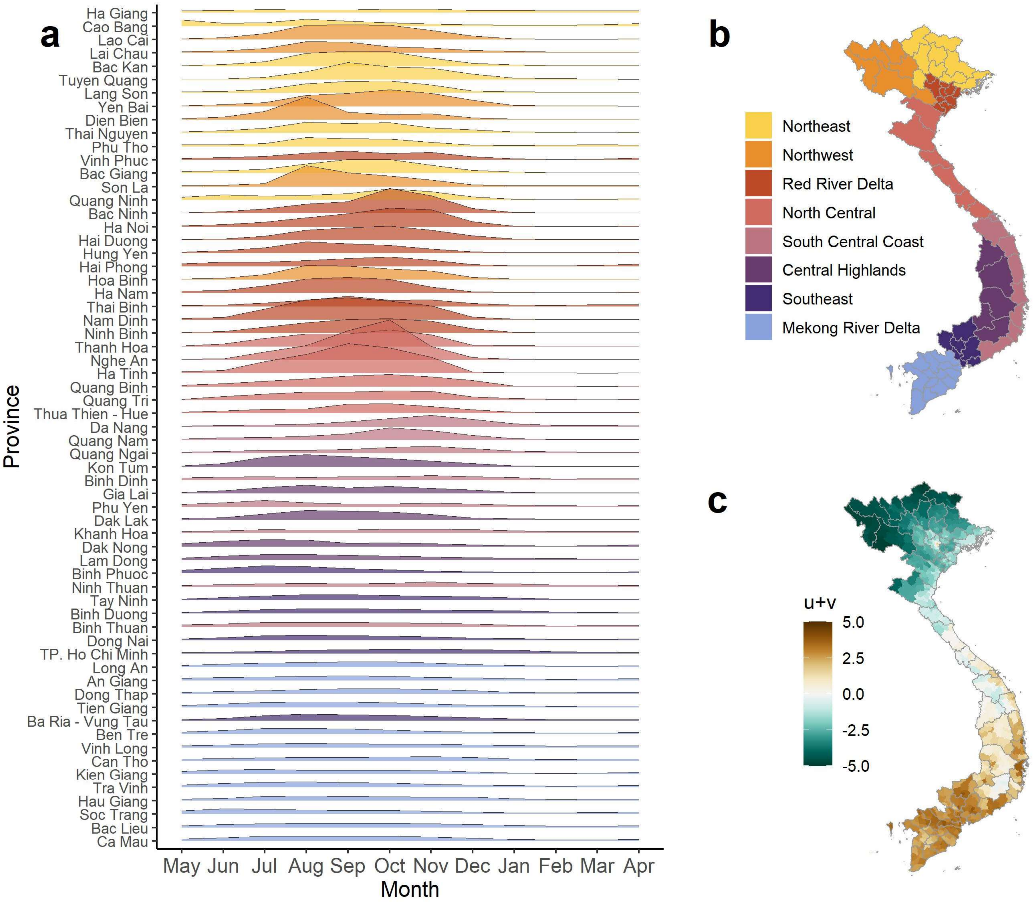

**Supp. Figure 6: Effect of individual covariates on model information criteria and unexplained random effects variation.** We selected the most appropriate functional form for each covariate by individually adding covariates to the baseline model. Graphs show changes in model information criteria (here, WAIC) and variation in spatiotemporal and seasonal random effects, measured as mean absolute error (Methods), when each covariate is added individually. Changes are shown relative to the baseline model (dashed line) and expressed as ΔWAIC – where more negative values indicate a greater improvement – and % change in random effects variation – where more negative values indicate that the covariate is explaining more of the unexplained variation. Point colour denotes covariate class (demography, mobility, urbanisation, infrastructure, temperature and hydrometeorology). We considered linear, logarithmic and nonlinear (random walk) terms for all covariates; only the best-fitting term is shown here for each. "Temporally-fixed" indicates that the model was fitted with the covariate held constant at its mean throughout the study period.

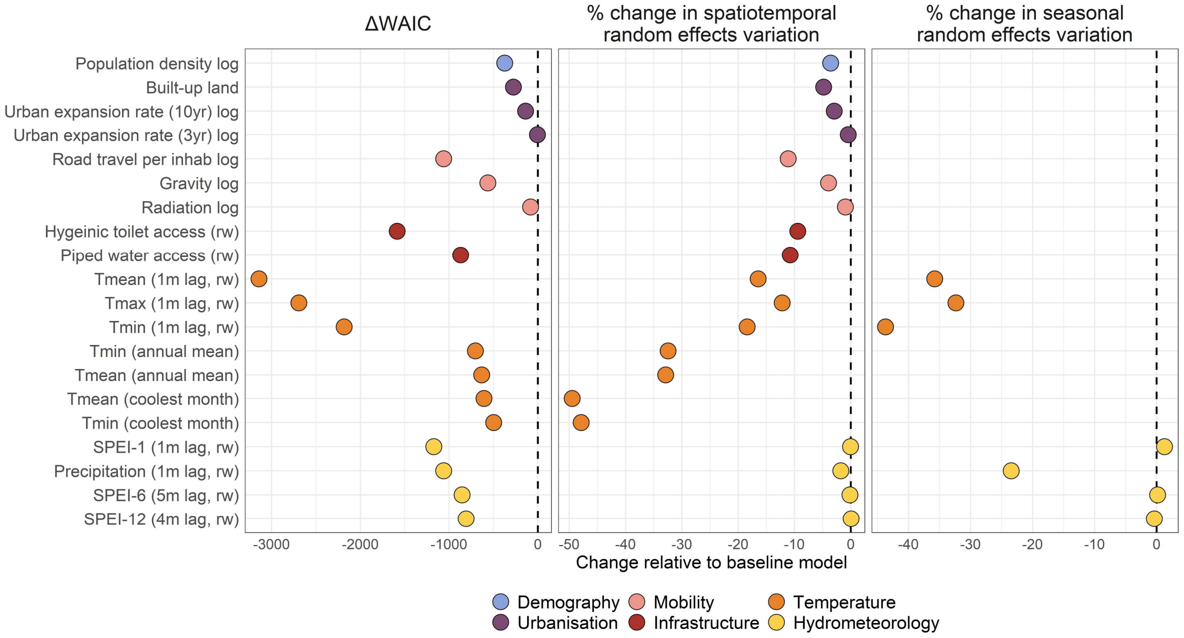

**Supp. Figure 7: Thermal suitability constraints explain the geographical gradient of dengue incidence across Vietnam.** We compared among 3 annual temperature metrics (annual mean $T_{mean}$, mean $T_{min}$, and $T_{mean}$ of the coolest month) in univariate analysis (Supp. Figure 6). The effect of adding each covariate individually on model WAIC (A) and district-level random effects variation ($MAE_{RE}$; B) are shown relative to the baseline model (dotted line). (C) Density distributions of fitted district-level random effect values (posterior mean) are shown for models with each covariate included individually (pink distribution), overlaid onto the distribution from the baseline model (C). $T_{mean}$ of the coolest month substantially reduced WAIC (A) and reduced the unexplained variation in spatiotemporal random effects by 50% (B). Maps show district-level random effects for baseline (D) and $T_{mean}$ coolest month (E) models (colour scale denotes per-district contribution to expected log dengue incidence, averaged across all years). Random effects magnitude reduces towards 0 in almost all locations when $T_{mean}$ coolest month is included (E), and shows that dengue incidence is higher than expected given temperature conditions in Ha Noi and much of south central coastal Vietnam (brown areas).

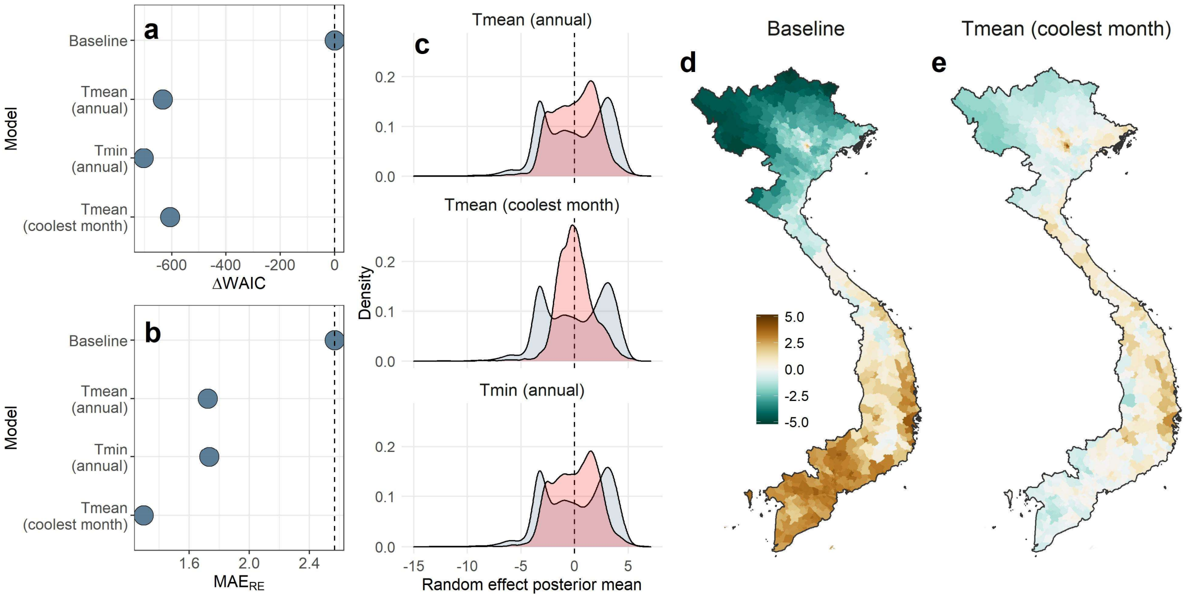

**Supp. Figure 8: Block cross-validation designs for predictive tests.** We used 3 block cross-validation designs to evaluate the contribution of covariates to out-of-sample prediction of dengue cases. Top panels (A-B) shows block designs for 10 example districts (10 rows) over 6 years of monthly observations. The dataset is randomly partitioned into 5 k-folds (A) according to either spatial (entire districts), spatiotemporal (district-year combinations) or seasonal block designs (within each district, 3-month blocks representing quarters of the year). Prediction error across all observations is calculated using predictions from 5 submodels with an 80-20 split into training (orange) and test (black) groups, with an example shown in B (here with k=1 as holdout group). An example of spatial and spatiotemporal holdout designs in the Mekong River Delta is shown in (C). Nearby locations provide information about expected dengue incidence in unobserved holdout districts (shown in red) via inferred spatial random effects (right-hand column), so these designs test which covariates help to predict observed departures from this expectation – i.e. spatial heterogeneity in incidence among nearby locations. Similarly, for seasonal blocks (D; top row), information about expected incidence during unobserved 3-month blocks (holdouts shaded pink) is contained within inferred annual district-specific and monthly random effects (D; bottom row). This design therefore tests which covariates improve prediction of monthly dynamics in a given year, i.e. reduce the gap between expected cases from the baseline random effects (red points) and actually observed cases (black points).

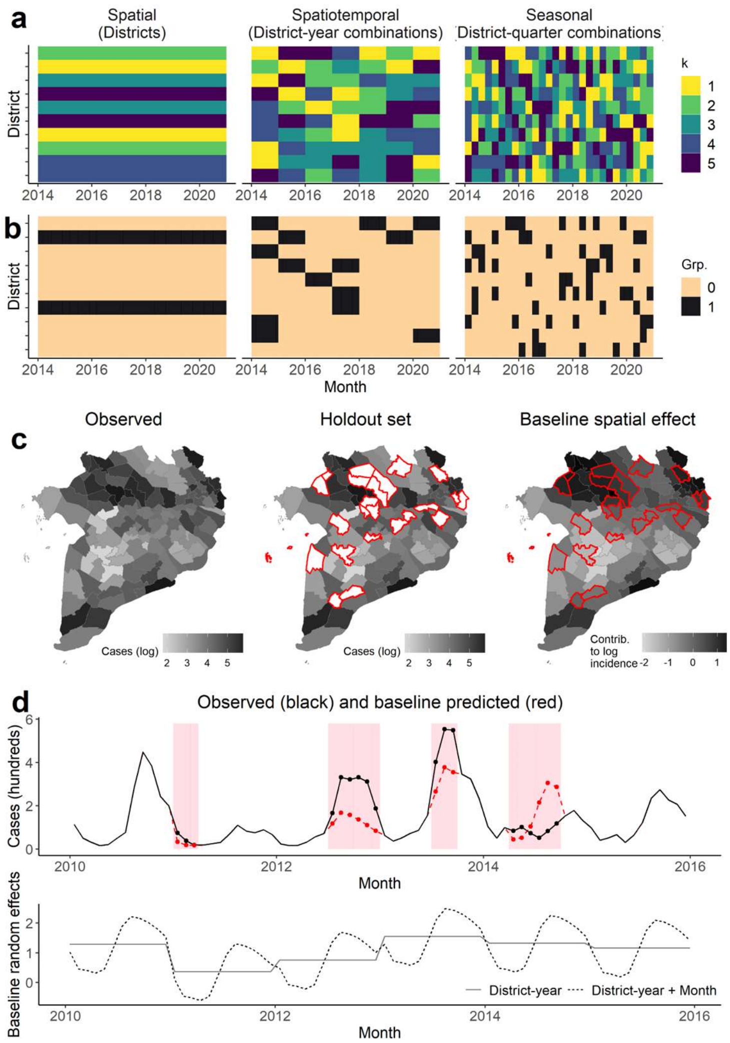

**Supp. Figure 9: Geographical changes in out-of-sample predictive error across Vietnam between baseline and full models under different block cross-validation designs.** Maps show, for each district, the change in mean absolute error between a baseline model (random effects-only) and the full multivariate model (including all covariates), calculated across all observations from 10 cross-validation repeats in each block holdout design. Maps are shown for 3 different block cross-validation designs, either spatiotemporal (holding out 20% of district-year combinations), spatial (holding out 20% of districts) or seasonal (holding out 20% of quarters within each district) (Supp. Figure 7). Colour scale denotes either a reduction (negative values; green) or an increase (positive values; brown) in mean out-of-sample prediction error in the full model relative to the baseline model, i.e. areas in green show where the full model improves over the baseline model, and vice versa. Nationwide summary metrics are shown in Figure 4.

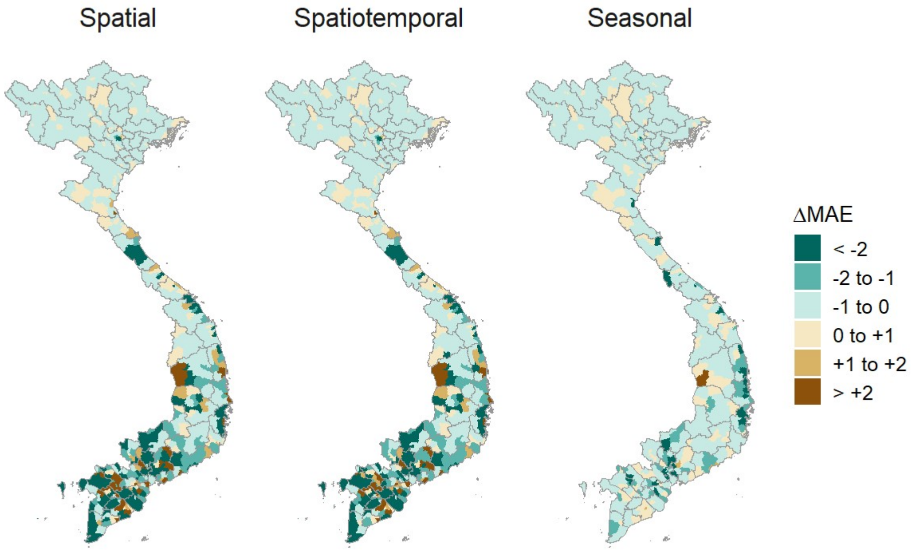

**Supp. Figure 10: Multivariate model sensitivity analysis by level of urbanisation.** Despite an overall lack of multicollinearity among covariates, highly urbanised areas (i.e. major and regional cities) tend to cluster with high values for several key variables. To ensure these are not biasing inferred relationships, we conducted a sensitivity analysis in which we fitted the full model (Figure 3) while excluding all observations from above each of 3 thresholds of urbanisation, defined as either more than 90% (excluding n=14,089 i.e. 8.1% of all observations), 70% (n=23,815; 13.6%) or 50% (n=33,760; 19.3%) of the population residing in urban areas according to the Vietnam Population and Housing Census (Methods). Models with lower thresholds are thus fitted to data from increasingly rural areas. Linear fixed effects (A; posterior marginal median and 95% credible interval) are shown on the link (logarithmic) scale, and nonlinear relationships (B-F) are shown on the relative risk scale, and point and line colour denotes urban holdout threshold. The magnitude, directional and shape of all relationships was robust to this sensitivity test.

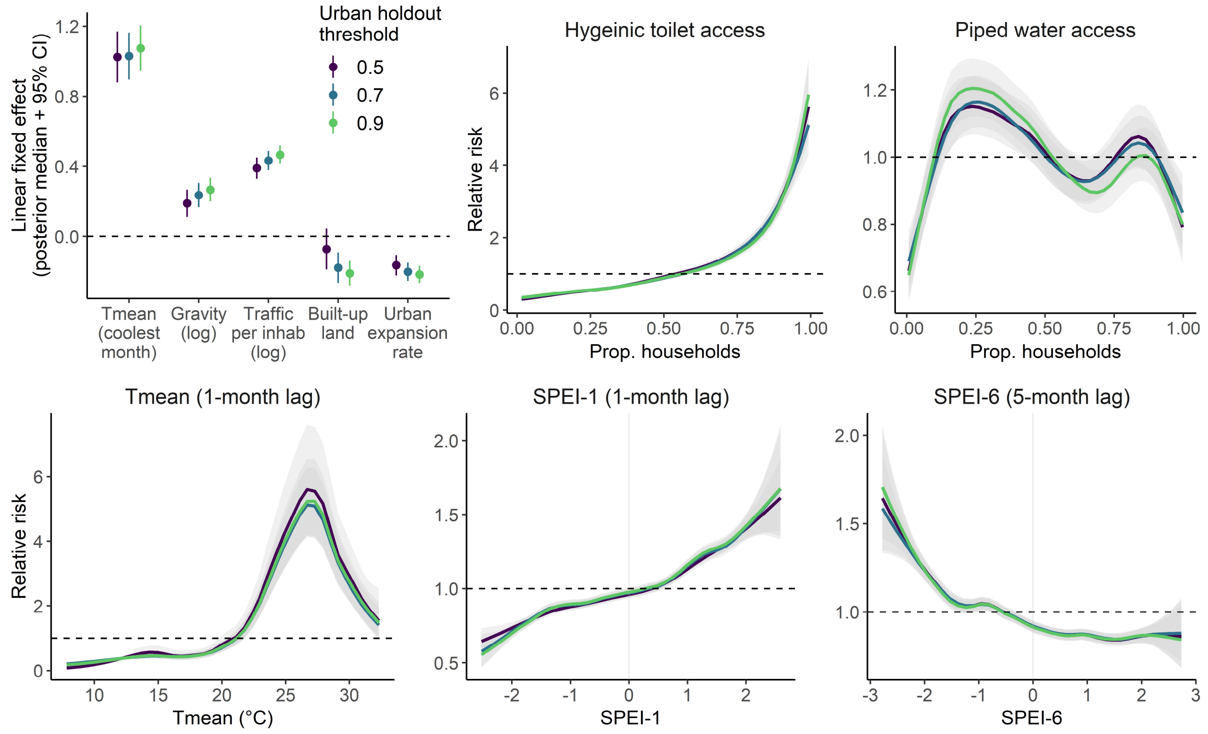

**Supp. Figure 11: Socio-environmental and climatic effects from region-specific models in southern and northern Vietnam.** Panels show posterior marginal intercept and linear fixed effects parameters (A-B; posterior median and 95% credible interval) and nonlinear effects specified as second-order random walks (C-G; posterior median and 95% credible interval) from 2 separate multivariate dengue incidence models, fitted to data from each broad subregion of Vietnam. Point or line colour denotes region as mapped in (H), either southern where dengue is present year-round (Mekong River Delta, Southeast, South Central Coast and Central Highlands, n=321 districts, 79,740 observations), or northern where dengue outbreaks are more sporadic (North Central, Red Diver Delta, Northeast and Northwest; n=346 districts, 95,196 observations). Intercept and fixed effect parameters are shown on the link scale (i.e. effects on log dengue incidence), and nonlinear effects are shown on the relative risk (exponentiated) scale.

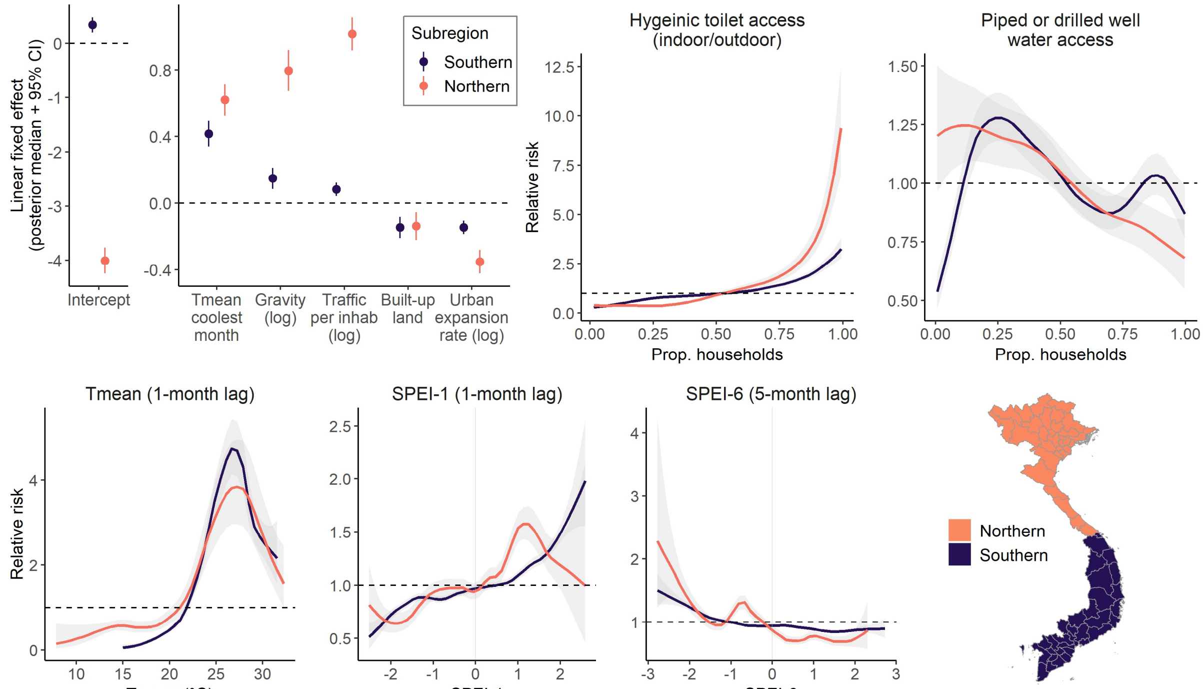

**Supp. Figure 12: Predictive influence of socio-environmental and climatic factors for subsets of northern and southern Vietnam.** To further examine differences in variable influence between southern (endemic) and northern (emerging) regions, we subset the observation-level predictions from the block cross-validation models (Figure 4) to calculate differences in out of sample mean absolute error for only northern (top row) and southern (bottom row) districts. Panels show spatial, spatiotemporal and seasonal block designs, and dandidate models excluding one covariate at a time from the full model are shown on the y axis, with the baseline (random effects-only) model for comparison. Point colour denotes broad covariate class: socio-environmental (green), climatic (blue) or baseline model (grey). Black points and error bars summarise the mean and 95% confidence interval across all 10 repeats. Values above zero indicate an increase in prediction error relative to the full model when a covariate is excluded (i.e. positive influence on prediction accuracy), and vice versa. Predictors are ordered on the Y-axis by their relative predictive influence in the spatial model; the top ranked predictors are mobility and temperature in the north, and infrastructure, urbanization and temperature in the south.

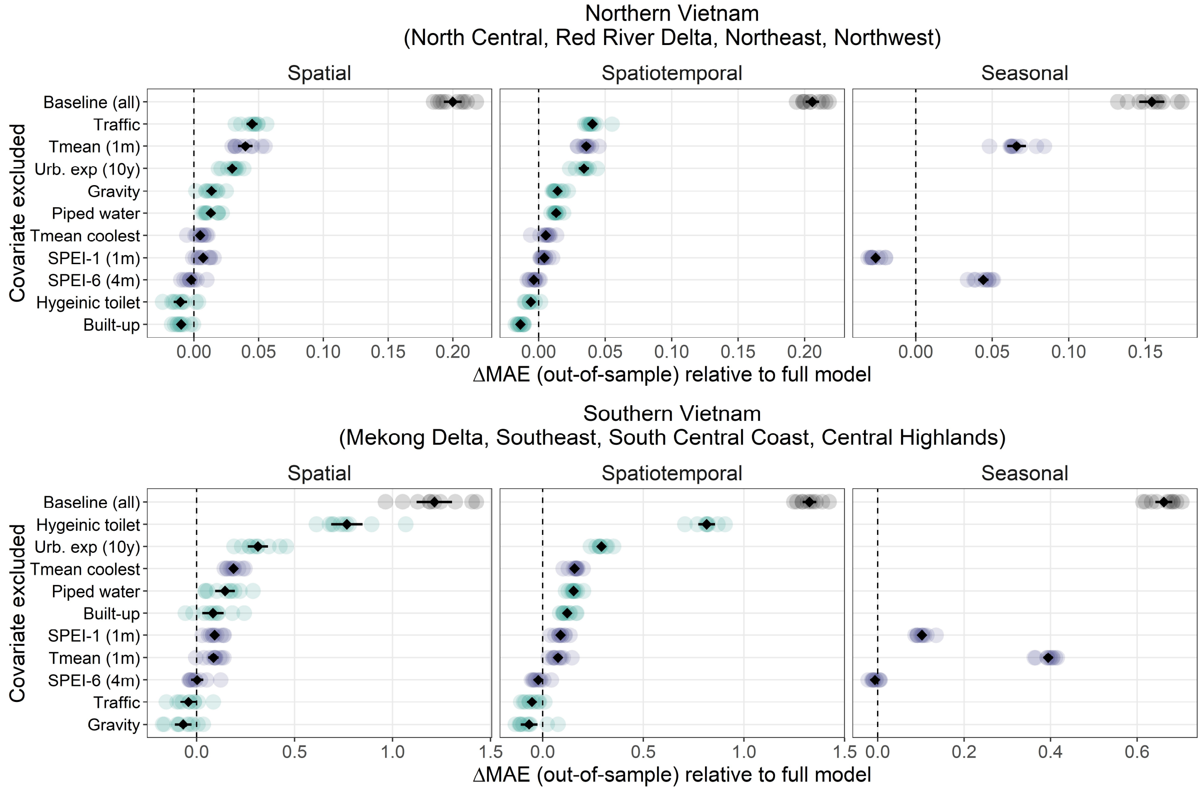

**Supp. Figure 13: Seasonal dengue relative risk dynamics between historical reference and present-day periods, for a subset of high change districts.** Graphs show long-term seasonal dynamics of temperature-driven dengue risk by dengue month (May to April) for 20-year reference period (1951-1970) and present day period (2001-2020). Individual years are shown as fine lines, and points, thick lines and error-bars show monthly 20-year mean and standard error, lines coloured by time period (green for reference period; purple for present-day). The top panel shows the top-ranked districts showing the greatest relative increase in risk for any single month (up to 56%; mainly from north and central highlands regions), and the bottom panel shows the top-ranked districts for the greatest relative decrease in risk for any single month (mainly from north central and Mekong Delta regions).

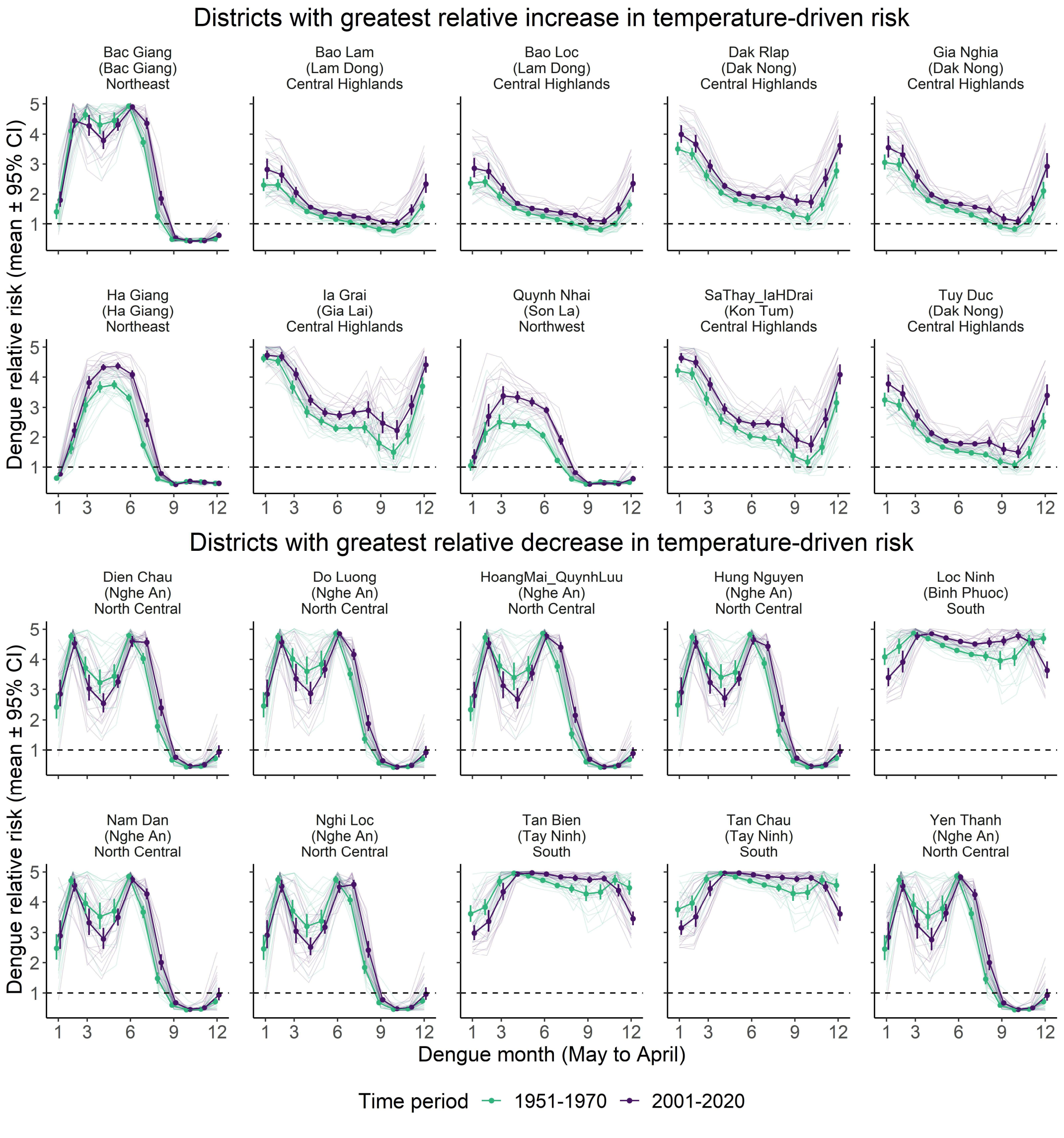

**Supp. Figure 14: Trends in seasonal monthly temperature patterns since 1950 from ERA5-Land, for high dengue burden cities.** Graphs show long-term seasonal dynamics of monthly average temperature ($T_{mean}$) by dengue month (May to April) between 1950 and 2020, for 5 cities with high dengue burden (main text, Figure 5). Individual years are shown as lines coloured by year (from pale green for 1950 to dark purple for 2020).

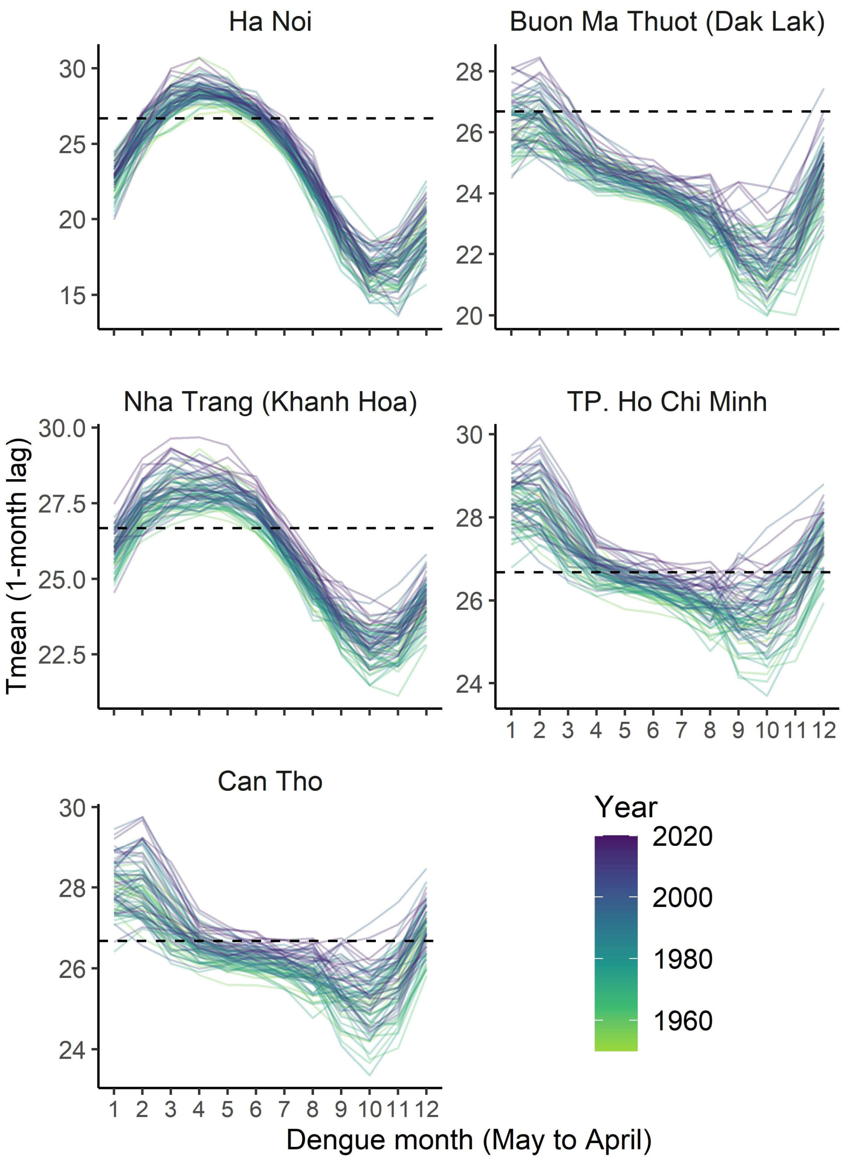

**Supp. Figure 15: Changes in temperature-driven risk from a 1971-1990 reference period.** As a sensitivity check, we repeated the projections of changing temperature-driven dengue risk (Figure 5) based on ERA5-Land temperature data from a later reference period (1971-1990) for which an order of magnitude more climate observation data is assimilated into the reanalysis[1]. Maps how the monthly percentage difference in 20-year average dengue risk between historical reference (1971-1990)  and present-day, with shading denoting directionality and strength of change (red denotes increasing risk and blue decreasing risk). Only statistically significant differences ($p<0.05$) are shown, with non-significant differences shaded white. Graphs show long-term dynamics and changes of seasonal risk by dengue month (May to April) for 5 cities with high dengue burden (Ha Noi in north; Buon Ma Thuot in the central highlands; Nha Trang on the south central coast; and Ho Chi Minh and Can Tho in the south). Individual years are shown as fine lines, and points, thick lines and error-bars show monthly 20-year mean and standard error, lines coloured by time period (green for reference period; purple for present-day).

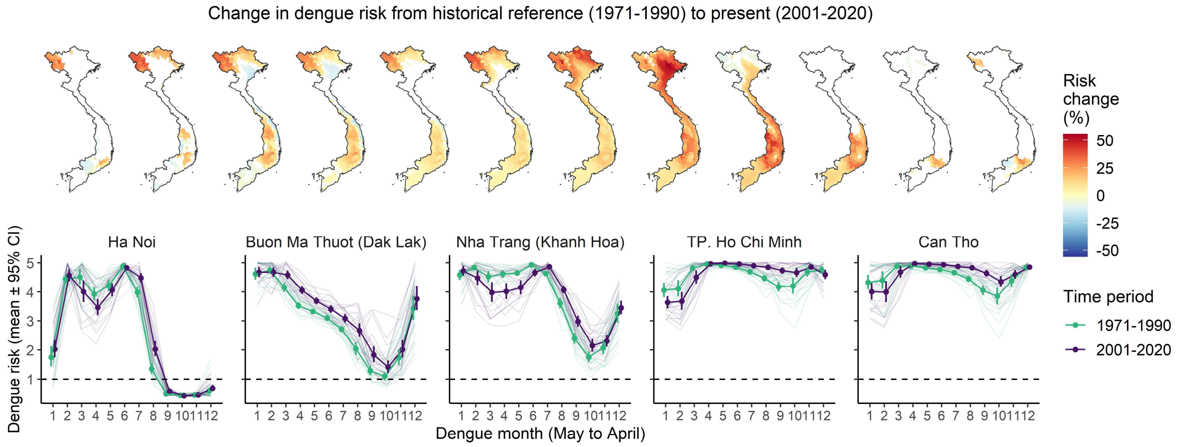

**Supp. Figure 16: Delayed and multiscalar effects of hydrometeorological extremes on dengue incidence.** Lines and ribbons show posterior marginal effects of Standardised Precipitation Evapotranspiration Index (mean and 95% credible interval) on dengue relative risk in southern Vietnam (n=321 districts, 79,740 observations). Rows denote SPEI at either short-timescale (SPEI-1) or long-timescale (SPEI-6), which measure standardised water surplus (>0) or deficit (<0) relative to the historical expectation for that locality (Methods). Columns denote temporal delay, from 0 to 6 months prior to focal month. Panels show 14 separate models, each adding an individual timescale-lag to the full model (Methods). Colour scale shows change in WAIC provided by adding each timescale-lag combination, with more negative values (darker colours) indicating greater improvement. Background panel shading denotes intensity of hydrometeorological extremes: near-normal to moderate (dark grey; 75.4% of observations for SPEI-1, 62% SPEI-6), moderate to severe (light grey; 24.6% SPEI-1, 38% SPEI-6) or extreme (0.8% SPEI-1, 3.1% SPEI-6).

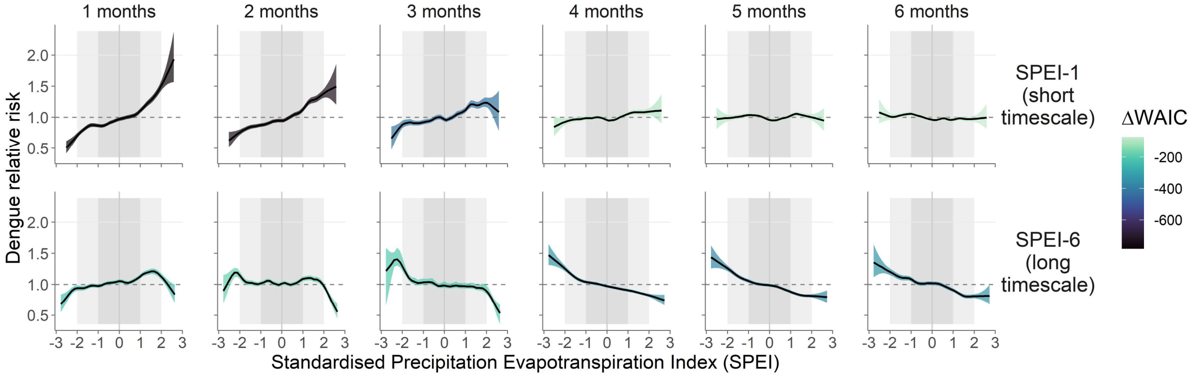

**Supp. Figure 17: Effects of interactions between drought metrics, improved water access and urbanisation on model information criteria for southern Vietnam.** We used the full multivariate model in southern Vietnam (including all socio-environmental covariates; Methods, Supp. Figure 11) to test whether the model was improved by including interactions between drought indicators (SPEI-1 1m and SPEI-6 5m lag) and either improved water access or urbanisation. Graph shows the difference in each of 3 information criteria (WAIC, DIC and cross-validated log score) between each candidate model and the full multivariate model with no interactions, where more negative values denote a greater improvement, and vice versa. For each SPEI metric we tested the effects of exclusion, interaction with water access, or interaction with urbanisation (low, medium or high categories; see Methods). Interactions between SPEI-6 and water substantially improved the model across all criteria, followed by slight improvements by including SPEI-6 and urban interaction, and no improvement of interactions with SPEI-1.

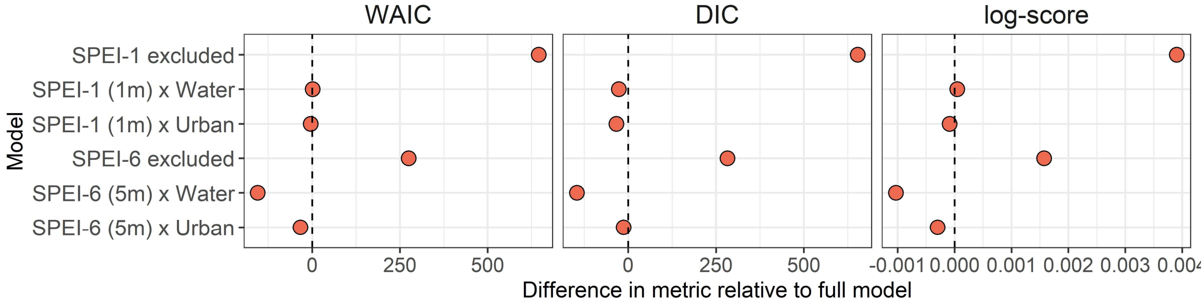

**Supp. Figure 18: Interactions between long-term drought and improved water access improve out-of-sample predictions of dengue incidence across southern Vietnam.** Figures show comparisons of out-of-sample prediction error between a full multivariate model (including all covariates and no interactions) and models either excluding SPEI metrics, or interacting SPEI with piped water access (Figure 6). Prediction error was calculated as mean absolute error (MAE) under 2 block cross validation designs, spatiotemporal and seasonal (Supp. Figure 8). Top row shows overall change in MAE between the full model (dashed line) and candidate models – where negative values denote a reduction in error, i.e. improvement – with red points and bars summarising the mean and 95% confidence interval across 10 repeats (blue points) (A). Middle row shows the percentage of districts (n=321) in which the candidate model reduces MAE compared to the full model (B), where higher values indicate a greater geographical extent of improvements (e.g., prediction error is reduced in 64% of districts when an interaction between SPEI-6 and water access is added to full model). Bottom row shows the geography of district-level changes in MAE from including an interaction between SPEI-6 and piped/drilled well water access. Colour scale denotes either a reduction (green shades) or an increase (brown shades) in MAE, averaged across all 10 repeats, compared to the full model without interaction.

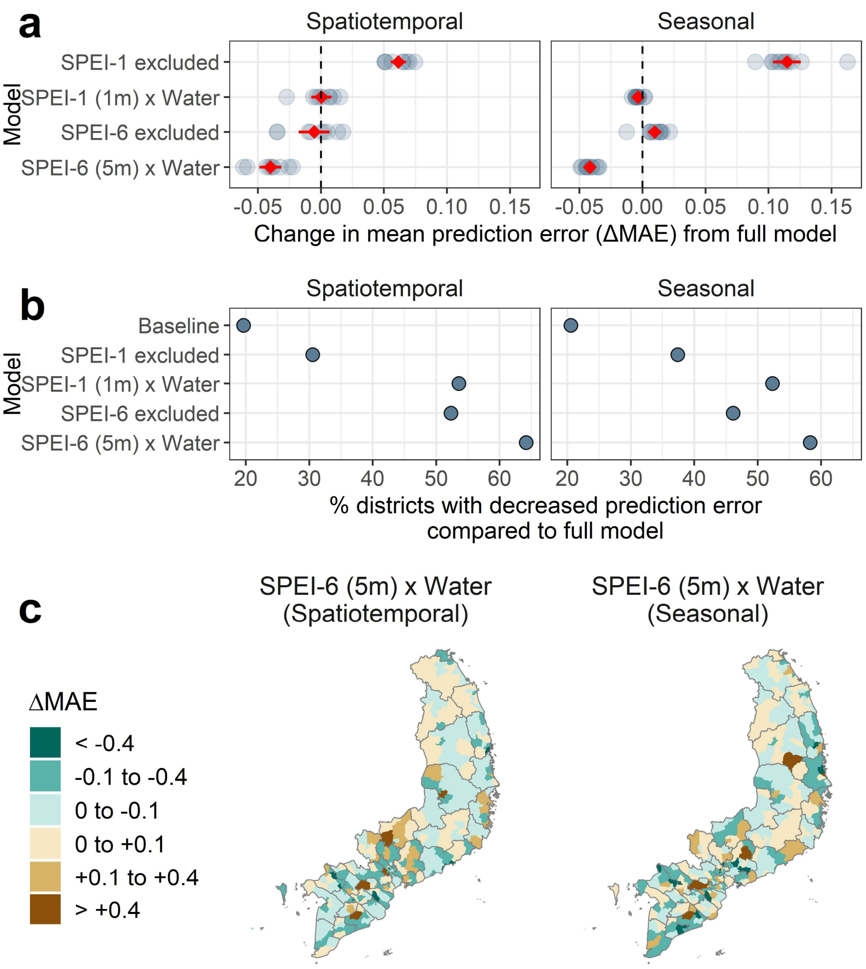

### Supp. Table 1: Description of covariates and associated data sources used in this study.

| Driver type | Covariate | Description | Data source | Temporal resolution |
|---|---|---|---|---|
| Demographic | Population density | Human population density (inhabitants/km$^2$) | CIESIN (2000), Vietnam Population and Housing Census (2009, 2019) | Annual (interpolated between observations) |
| Urbanisation | Built-up land | Population-weighted built-up land area, identified using satellite land cover data and weighted by WorldPop population layer. | ESA-CCI Land Cover (300m) | Annual |
| Urbanisation | Urban land expansion rate (3year and 10year windows) | Expansion rate of impervious (built-up) land area in the preceding 3 year and 10 year windows (km$^2$/year) | Landsat-derived 30m annual impervious surface (Liu et al 2020). | Annual |
| Infrastructure | Hygienic toilet access | Proportion of district households with access to an indoor or outdoor hygienic (flush) toilet | Vietnam Population and Housing Census (2009, 2019) | Annual (interpolated between observations) |
| Infrastructure | Piped water access | Proportion of district households with access to either piped tap or drilled well water | Vietnam Population and Housing Census (2009, 2019) | Annual (interpolated between observations) |
| Mobility | Road traffic mobility | Province-level annual estimates of road traffic mobility (mean kilometres travelled per inhabitant). | Vietnam General Statistics Office | Annual |
| Mobility | Connectivity | Mean of pairwise potential population fluxes between focal district and all other districts, predicted by general human mobility models (gravity and radiation). | Gravity and radiation models applied to district-level annual population counts. | Annual |
| Climate | Air temperature annual | Annual mean Tmean and mean Tmin (across all months) and Tmin of the coolest month | ERA5-Land (via Copernicus Climate Data Service) | Annual |
| Climate | Air temperature monthly (Tmean, Tmin, Tmax) | Monthly average of daily mean, minimum and maximum air temperature, derived from hourly gridded air temperature estimates from ERA5-Land reanalysis. | ERA5-Land (via Copernicus Climate Data Service) | Monthly, at 0 to 6 month lags |
| Climate | Precipitation | Mean daily precipitation (mm) derived from hourly gridded WFDE5 precipitation reanalysis data (ERA5 bias corrected using CRU and GPCC precipitation datasets). | WFDE5 (via Copernicus Climate Data Service) | Monthly, at 0 to 6 month lags |
| Climate | Standardised Precipitation Evapotranspiration Index (SPEI) | Multiscalar drought indictor calculated across 1, 6, and 12 month windows to represent different drought timescales. | Derived from WFDE5 precipitation and ERA5 evapotranspiration data using R package 'spei' | Monthly, at 0 to 6 month lags |

**Supp. Table 2: Summary metrics for baseline and full multivariate models of dengue incidence.** Table shows, for the baseline and full models of dengue incidence, summary metrics Deviance Information Criterion (DIC), Watanabe-Akaike Information Criterion (WAIC), cross-validated logarithmic score (log-score) and mean absolute error in spatiotemporal district-level random effects ($MAE_{RE}$). Bottom row shows the difference in the metrics between the baseline and full models, with more negative values denoting greater improvement in all metrics.

| Model | DIC | WAIC | log-score | $MAE_{RE}$ |
|---|---|---|---|---|
| Baseline | 606149.1 | 607669.1 | 1.769 | 2.567 |
| Full | 598797.6 | 600910.8 | 1.745 | 1.353 |
| Difference | -7351.6 | -6758.3 | -0.024 | -1.214 |

**Supp. Table 3: Parameter and hyperparameter estimates from the full multivariate model of dengue incidence.** Tables show posterior marginal fixed effects parameter and hyperparameter estimates (median and 95% credible interval) from the full model of dengue incidence. Incidence was modelled using a negative binomial error distribution with log-link, so estimates are on the natural logarithmic scale. Fixed effect covariates were log transformed (where specified) or otherwise were scaled (mean 0, sd 1) prior to model fitting so parameter estimates reflect the effect of a change in 1 scaled unit (standard deviation) of the covariate on the response variable (Methods).

| Parameter | Type | Median | CI_0.025 | CI_0.975 |
|---|---|---|---|---|
| Intercept | Fixed effect | -1.843 | -2.082 | -1.607 |
| Tmean coolest month | Fixed effect | 1.124 | 1.005 | 1.244 |
| Gravity (log) | Fixed effect | 0.287 | 0.227 | 0.348 |
| Built-up land | Fixed effect | -0.143 | -0.195 | -0.092 |
| Urban expansion rate (10 year, log) | Fixed effect | -0.251 | -0.288 | -0.213 |
| Traffic per inhab (log) | Fixed effect | 0.459 | 0.41 | 0.508 |
| Size (1/overdisp) for neg. binom | Hyperparameter | 1.592 | 1.573 | 1.616 |
| Precision for Month (RW1) | Hyperparameter | 3.969 | 3.534 | 4.645 |
| Precision for District (BYM2) | Hyperparameter | 0.13 | 0.124 | 0.138 |
| Phi for District (BYM2) | Hyperparameter | 0.905 | 0.894 | 0.914 |
| Precision for Hygienic toilet (RW2) | Hyperparameter | 23.834 | 11.224 | 57.789 |
| Precision for Piped water (RW2) | Hyperparameter | 14.604 | 7.31 | 29.163 |
| Precision for Tmean 1m (RW2) | Hyperparameter | 5.719 | 4.01 | 7.661 |
| Precision for SPEI-1 1m (RW2) | Hyperparameter | 35.582 | 20.53 | 65.605 |
| Precision for SPEI-6 5m (RW2) | Hyperparameter | 18.973 | 9.272 | 44.47 |

**Supp. Text 1: Data sources and processing of socio-environmental and climatic covariates**

Population and population density

District-level population counts were accessed for three time points: 2000, 2009 and 2019. Population for 2000 was extracted from Gridded Population of the World v4 (https://sedac.ciesin.columbia.edu/data/collection/gpw-v4) because this dataset was produced using census observations from 1999, and census results were not available in report form for this year. For 2009 and 2019, population counts and urban population counts were obtained directly from the Vietnam Population and Housing Census reports for the respective years (accessed via the Vietnam General Statistics Office, VGSO; https://www.gso.gov.vn/en/homepage/). To produce annual estimates of population, in each district we linearly interpolated between time points, and back-projected to 1998 and forward-projected to 2020 assuming the same annual growth rate as in the subsequent or preceding 10 years. Population density at each time point was calculated as population divided by district area.

Temperature (monthly)

We derived temperature metrics by processing hourly gridded air temperature layers from the high-resolution ERA5-Land reanalysis dataset[2,3] (~9km resolution), accessed through the Copernicus Climate Data Service (CDS; https://cds.climate.copernicus.eu/). Hourly rasters were aggregated to create daily rasters of per-grid cell mean temperature ($T_{mean}$), minimum temperature ($T_{min}$) and maximum temperature ($T_{max}$). All daily rasters for each month were then aggregated to calculate per-grid cell monthly means of daily $T_{mean}$, $T_{min}$ and $T_{max}$. We then extracted mean monthly $T_{mean}$, $T_{min}$ and $T_{max}$ across all non-NA grid cells within each district polygon (n=667) using 'exactextractr'[4]. If all grid cells in a district were NA (for some offshore island and small coastal peninsula districts), the district was assigned the same climate time series as its nearest neighbour. To represent seasonal and delayed effects of air temperature on dengue transmission, for each district we calculated air temperature metrics in a 2-month window starting at a lag of 0 to 6 months prior to each focal month (for example, *"$T_{mean}$ 1m"* is average $T_{mean}$ across 1 to 2 months prior to focal month).

Temperature (annual)

Broad gradients in thermal suitability could confound the relationship of dengue with other covariates that also vary geographically across Vietnam (e.g. mobility and piped water access are generally lower in higher altitude areas; Supp. Figure 2). To account for this, we also calculated 3 annual bioclimatic indicators to represent more fundamental constraints on dengue establishment and persistence, using monthly observations extracted from ERA5-Land as described above. For each district and year we calculated mean $T_{mean}$ across all months (*$T_{mean}$ annual*), mean $T_{min}$ across all months (*$T_{min}$ annual*), and $T_{mean}$ of the coolest month of the year (*$T_{mean}$ coolest month*) (Table 1). $T_{mean}$ annual and $T_{min}$ annual reflect average suitability for dengue transmission across a year (daily mean temperature, and daily minimum temperature respectively). $T_{mean}$ coolest month reflects that temperature constraints on *Aedes* survival, reproduction and vector competence are likely greatest during the cooler months, which could affect the potential for dengue to persist year-round in some areas (for example, high altitude and latitude regions where intra-annual temperature ranges are widest; Supp. Figure 3).

Precipitation (monthly)

We derived precipitation metrics by processing hourly ERA-based gridded reanalysis products. Since precipitation can vary widely over quite small geographical areas, we compared data products against monthly weather station from Vietnam (2002 to 2019) to inform choice of  the most accurate dataset. We used WFDE5 v2.1 precipitation data (ERA5 precipitation bias-corrected with reference to CRU and GPCC gridded weather station observations[5]) during available years (up to 2019), as the use of bias-corrected precipitation is generally recommended for reanalysis data, and these data proved substantially more accurate when benchmarked against weather station data (Supp. Figure 4). From 2020 onward (Jan 2020 to April 2021) we used ERA5-Land data because bias-corrected data were not yet available for these years. As with temperature, we aggregated hourly rasters to calculate daily precipitation, then aggregated to month by calculating monthly mean daily precipitation. We extracted monthly precipitation for each district polygon, and calculated mean in 2 month windows at lags of 0 to 6 months, as described above for temperature.

Standardised Precipitation Evapotranspiration Index (monthly)

To evaluate multiscalar effects of hydrometeorological extremes, we derived district-level monthly time series of Standardised Precipitation Evapotranspiration Index (SPEI) from 40-years of bias-adjusted precipitation (WFDE5 1981-2019; ERA5-Land 2020-2021) and potential evapotranspiration data (ERA5-Land 1981-2020) derived from ERA5 reanalysis. SPEI is a multiscalar drought indicator that incorporates effects of both precipitation and temperature (via evapotranspiration) on water availability, and measures hydrological deficit (values below 0) or excess (values above 0) in a particular time window relative to the long-term historical average for the same period (for example, a 6-month SPEI for Jan-Jun 2018 would compare to Jan-Jun in all other years)[6,7]. Different time windows of SPEI represent varying timescales of hydrometeorological excess or deficit: transient excess or deficit (in a 1 month window i.e. SPEI-1) may affect patterns of surface and soil water, whereas medium-to- long-term excess or deficit may affect water availability from reservoirs (SPEI-6) or groundwater storage (SPEI-12 or longer)[7]. For each district we extracted 40-year time series of monthly total precipitation and potential evapotranspiration from data sources as described above. We then used the R-package 'spei'[8] to derive time series of monthly SPEI, calculated in time windows of the preceding 1 (SPEI-1), 6 (SPEI-6) and 12 months (SPEI-12) (Supp. Figure 3). As with temperature and precipitation, we then calculated the 2-month window mean for each metric, at lags from 0 to 6 months. SPEI-1 was not correlated to SPEI-6 and SPEI-12 ($\rho$ of 0.55 and 0.42 respectively), but SPEI-6 and SPEI-12 were strongly correlated to one another ($\rho$ = 0.79).

Built-up land cover

We extracted annual district-level *built environment extent* from ESA-CCI land cover grids (300m resolution), which are available annually from 1992 to 2018, to represent exposure to preferential habitats for *Ae. Aegypti*. For each year we used 'exactextractr' to calculate the proportion of grid cells defined as urban (i.e. where urban is the dominant land cover within the 300m grid cell), weighted by population accessed from WorldPop annual unconstrained 100m population rasters (https://dx.doi.org/10.5258/SOTON/WP00645). We weighted by population to improve comparability across districts (i.e. to more closely reflect population exposure to built environments), since different districts vary in area, shape and population distribution. In each district we forward projected to provide estimates in missing years (2019-20) assuming the same urban change rate as in the previous 5 years.

Urban expansion rate

Economic transitions in the last 30 years have seen expansion of urban areas across much of Vietnam, particularly around major cities such as Ha Noi and Ho Chi Minh city, as well as regional hubs such as Dong Nai and Binh Duong. Rapid short-term expansion of urban environments is often hypothesised to be a general driver of increased dengue risk, because disturbed habitats with variable drainage quality (e.g. construction sites or informal settlements) often contain a particularly high density of suitable *Ae. aegypti* breeding habitat. Conversely, planned expansion of urban environments may, at longer timescales, lead to local improvements in housing quality, drainage and sanitation systems, and increases in vector control efforts, that reduce breeding site habitat and thus dengue risk. To test for associations between urban expansion rates and dengue risk at different timescales, we used very high-resolution (~30m) annual rasters of impervious surface for the period 1985 to 2015, derived from Landsat data[9]. For each district and year we calculated *urban expansion rates* ($km^2$/year) within the preceding 3 years (short-term growth) and 10 years (long-term growth). To produce estimates for missing years, we projected forward from 2016 to 2020 in each district assuming the same change rate as the previous 5 years. Short-term and long-term growth were highly correlated ($\rho$ = 0.84; Supp. Figure 4).

Household water supply and sanitation infrastructure

Since dengue transmission generally occurs in and around homes[10,11], quality and type of household water supply and sanitation infrastructure often correlates with individual-level dengue risk[12]. Less is known about their importance in shaping broad-scale spatiotemporal distributions of incidence, although there is increasing evidence for complex effects of household water access, where irregular or unreliable water supply may increase households' propensity to store water around homes (and thus create breeding sites for *Aedes* mosquitoes). From the 2009 and 2019 Vietnam Population and Housing Census, we obtained district-level estimates of proportion of households with access to hygienic toilet facilities (either indoor or outdoor flush toilet, as an indicator of overall sanitation quality) and piped or drilled well water (i.e. a relatively reliable clean water supply). 2009 data were accessed via the World Bank's mapVietnam portal (http://www5.worldbank.org/mapvietnam/) and 2019 data were accessed via the VGSO's geospatial data portal (https://gis.gso.gov.vn/). To obtain annual estimates in each district, we linearly interpolated between years, and back-projected in annual timesteps to 1998 assuming the same annual growth rate as throughout the 2009 to 2019 period. This assumption was consistent with national-level aggregate statistics (via VGSO) showing a year-on-year national-level increase in hygienic water and sanitation access during this early period.

Road travel

Human movement and geographical connectivity across both short and long-ranges are important drivers of dengue diffusion, outbreak dynamics and the reintroduction of dengue viruses[13–15]. To capture geographical and temporal differences in overall rates of movement, we accessed province-level annual data on total kilometres travelled by road, from the VGSO. Data were available at province level from 2000 to 2020. Time series for several provinces started in 2003, and were aggregated with other provinces before this year (Dak Nong combined with Dak Lak; Lai Chau with Dien Bien; Hau Giang with Can Tho). We disaggregated these pairs of provinces during earlier years by back-projecting those provinces from 2003 to 2000 assuming the same annual rate of change as in the subsequent 3 years, and then subtracting these estimates from the aggregated province in the years 2000 to 2002. For each province, we then back-projected to 1998 assuming the same annual rate of change as during 2000 to 2003. Finally, we adjusted for population by calculating the annual road travel distance per inhabitant, by dividing total kilometres travelled by the number of province inhabitants in that year. This covariate was included in models at province-level, as it was unfeasible to disaggregate further to district-level (Supp. Figure 2).

Gravity and radiation flux (connectivity)

Ideally the relative connectivity of districts (population fluxes) would be calculated using empirical data such as mobile phone records, however sufficiently long and internally-consistent time series are not available for our study period. We therefore use general human movement models[16] to predict annual connectivity (relative population flux), based on annual population estimates and a matrix of pairwise estimated travel times between all districts. Here, we use gravity and radiation models, both of which are based on the assumption that the rate of travel between a given pair of locations (here, districts) is proportional to their relative population sizes (i.e. more populated areas attract more travel) and declines with distance. The gravity model represents the economic "pull" of more populated centres, and the radiation model introduces an additional constraint that accounts for the competing attractiveness of other populated centres within a similar distance[17]. Gravity models have been shown to be useful for describing directed movements (e.g. commuting patterns into major cities) in both high- and low-mobility settings, whereas radiation models have often been shown to be more accurate at approximating movements among rural areas and smaller cities, although the performance of both varies across contexts[16].

We used a published friction raster and least-cost travel algorithm[18] to develop a matrix of pairwise, population-weighted travel times between all pairs of districts within Vietnam (i.e. the estimated mean travel time between any pair of persons across two districts), with WorldPop population 2015 used as a population weighting layer. We used travel time because Vietnam's wide altitudinal and latitudinal range means that simpler distance-based metrics may substantially underestimate the time taken to travel between certain pairs of locations.

We used a naïve (parameter-free) gravity model and radiation model to predict the pairwise relative flux between each pair of districts in each year. The naïve gravity model assumes that the potential gravity flux ($G_{ij}$) between two locations *i* and *j* is influenced only by the populations of the source and destination locations and the intervening distance, and is defined as in Wesolowski et al. (2015)[13]:

$$G_{ij} = \frac{p_i p_j}{d_{ij}}$$

where $p_i$ and $p_j$ are the populations of source *i* and destination *j* respectively, and $d_{ij}$ is estimated travel time between districts *i* and *j*.

Potential radiation flux ($R_{ij}$) between two districts is predicted as defined in Simini et al. (2012)[17]:

$$R_{ij} = T_i \frac{p_i p_j}{(p_i + s_{ij})(p_i + p_j + s_{ij})}$$

where $p_i$ and $p_j$ are as above, $T_i$ is the number of inhabitants that start their journey from source location *i* (which is proportional to the source population, so here is set to equal $p_i$)[17], and $s_{ij}$ is the total population within the travel time radius of $d_{ij}$ (i.e. residing within the same or lower travel time as destination *j*), excluding the source and destination populations.

Finally, the annual pairwise matrices of flux estimates were used to calculate annual summary metrics of connectivity: mean district-level *gravity flux* and *radiation flux* (i.e. mean predicted flux between the focal district and all other districts in the same year).
